# Genetically Predicted IL-18 Inhibition and Risk of Cardiovascular Events: A Mendelian Randomization Study

**DOI:** 10.1101/2024.07.01.24309808

**Authors:** Stephen O. Brennan, Peter J. Kelly, Sarah Gorey, Pádraig Synnott, Dipender Gill, Martin Dichgans, Marios K. Georgakis, Marie-Joe Dib, Eloi Gagnon, Niall Mahon, Gavin J. Blake, Christina Jern, Hugh S. Markus, William Whiteley, John J. McCabe

## Abstract

**Background:** Inflammation is an emerging target for the prevention and treatment of cardiovascular disease (CVD). This drug-target Mendelian randomization (MR) study aimed to predict the on-target effects of IL-18 inhibition on CVD risk. Furthermore, we aimed to explore the effects of IL-18 inhibition on cardio-metabolic traits, cardiac structure, and function, and identify potential adverse outcomes.

**Methods:** We selected five independent circulating IL-18-lowering variants around the *IL-18* gene locus from the Systematic and Combined AnaLysis of Olink Proteins (SCALLOP) consortium. We then performed two-sample MR analyses to investigate the association of genetically proxied IL-18-inhibition on downstream inflammatory markers, risk of CVD, cardiac magnetic resonance (CMR) imaging measurements of cardiac structure and function, cardiometabolic traits, and a selection of potential adverse effects. We utilized data from the UK Biobank, Cardiogram, GIGASTROKE, and other large genomic consortia (sample range: 3,301-1,320,016).

**Results:** Following correction for multiple comparisons, one standard deviation (SD) lower in genetically-predicted circulating IL-18 was associated with reductions in downstream biomarkers of IL-18 signaling, including C-reactive protein (SD change -0.02, 95% CI -0.03, -0.02), tumor necrosis factor (SD change -0.19, CI -0.25, -0.14), interferon-gamma (SD change -0.15, CI -0.22, -0.08), and CXCL10 (SD change -0.13, CI -0.16, -0.09). Lower genetically-predicted IL-18 levels were associated with reduced risk of cardioembolic stroke (Odds Ratio [OR] 0.85, CI 0.79-0.92), but not other stroke subtypes. Furthermore, lower genetically predicted IL-18 levels were associated with reduced risk of peripheral arterial disease (OR 0.91, CI 0.84-0.97), atrial fibrillation (OR 0.94, CI 0.89-0.99), and heart failure (OR 0.84, CI 0.77-0.92), as well as improvements in CMR traits, including a reduction in left atrial volume (β -0.02, CI -0.03, -0.00). Lower genetically-predicted IL-18 levels were associated with lower risk of chronic kidney disease, autoimmune diseases, a favorable cardio-metabolic profile, and higher odds of lung cancer, but not infections.

**Conclusions:** Our study provides genetic support that impaired IL-18 signaling may be causally associated with a lower risk of cardioembolic stroke, possibly mediated through prevention of cardiac re-modelling, heart failure and atrial fibrillation. IL-18 represents a potential target for anti-inflammatory therapy in stroke and CVD that warrants further investigation in clinical trials.

**Clinical Perspective:** What is new?

- Using multi-omic data, this Mendelian Randomization study provides evidence that IL-18 lowering is associated with a lower lifetime risk of cardiac remodeling, heart failure, and cardioembolic stroke.
- A significant proportion of the protective effect of impaired IL-18 signaling on cardioembolic stroke was mediated through a reduced risk of AF.

What are the clinical implications?

- These data provide compelling evidence that the IL-18 signaling pathway is a promising druggable target for the treatment of heart failure and the prevention of cardioembolic stroke.
- Several monoclonal antibodies targeting IL-18 are in development for the treatment of atopic dermatitis and could be considered for re-purposing for cardiovascular disease.

## Introduction

Inflammation has emerged as a promising target for the treatment of atherosclerosis and the secondary prevention of coronary events.^1,2^ The first anti-inflammatory trials for secondary prevention after stroke have primarily focused on the therapeutic potential of these agents in patients with atherosclerotic mechanisms and excluded patients with cardioembolic stroke.^3^ However, several complementary lines of evidence also support the importance of inflammatory mechanisms in the pathophysiology and progression of heart failure (HF), atrial fibrillation (AF), and cardioembolic stroke. Animal models provide compelling data that the NOD-like receptor family pyrin domain-containing 3 (NLRP) inflammasome is upregulated in cardiomyocytes in the pathogenesis of AF and promotes structural and electrical atrial remodeling.^4^ Inflammatory signaling is implicated in several pathways central to the pathophysiology of HF, including mitochondrial dysfunction, calcium homeostasis, cardiac fibrosis, and impaired cardiomyocyte contractility.^5,6^ Inflammatory markers are associated with incident AF in population-based studies^7,8^ and AF-related stroke.^9^ Despite these promising data, the results of the first RCTs of anti-inflammatory therapies in HF,^10,11^ and AF^12^ have been neutral.

The NLRP3 inflammasome mediates some of its effects through activation of interleukin-1β (IL-1β) via caspase-1 and leads to downstream IL-6 expression. Pre-clinical,^13^ observational,^14,15^ and genetic studies^16^ have coalesced to identify the IL-1β-interleukin-6 (IL-6) signaling pathway as a promising target for risk reduction in cardiovascular disease^17^ and stroke.^18^ Significantly less attention has been given to the role of interleukin-18 (IL-18), which is also cleaved to its active form by caspase-1 following activation by the NLRP3 inflammasome.^13^ IL-18 is a potent pro-inflammatory cytokine that promotes atherosclerotic plaque progression in pre-clinical work.^13^ However, experimental data have also demonstrated that IL-18 signaling plays a key role in cardiomyocyte hypertrophy, ventricular dysfunction, and cardiac fibrosis.^19–22^ Elevated IL-18 levels are observed in patients with HF and correlate with disease severity, but whether this inflammatory response is a cause or a consequence of the pathogenesis of HF is unknown.^23^ Some, but not all, prospective cohort studies have reported that IL-18 is associated with incident coronary events^24–26^ and HF,^24,27^ but not incident stroke.^24^ It remains uncertain whether pharmacological inhibition of the IL-18 signaling pathway is a viable treatment option for a variety of cardiovascular diseases, including coronary disease, stroke, AF, and HF.

In the absence of RCTs of therapeutics targeting the IL-18 signaling pathway for the treatment of cardiovascular disease, genetic studies can offer valuable insights into the on-target effect of these pharmacological agents.^28,29^ Drug-target Mendelian Randomization (MR) employs genetic variants associated with the levels or activity of drug-target proteins, thus serving as proxies for pharmacological agents.^29^ The primary aim of this study is to explore the potential benefit of IL-18 inhibition on cardiovascular outcomes using a drug-target MR approach.

## Methods

### Study design

We used MR to assess the causal association between genetically proxied downregulation of IL-18 signaling and: 1) cardiovascular disease outcomes, with replication in non-overlapping samples where possible; 2) cardiometabolic and inflammatory traits, including cardiac magnetic resonance (CMR) measures of cardiac structure and function; and 3) potential adverse and beneficial effects associated with reduced IL-18 signaling. An overview of the study design is detailed in Figure 1.

**Figure 1.**
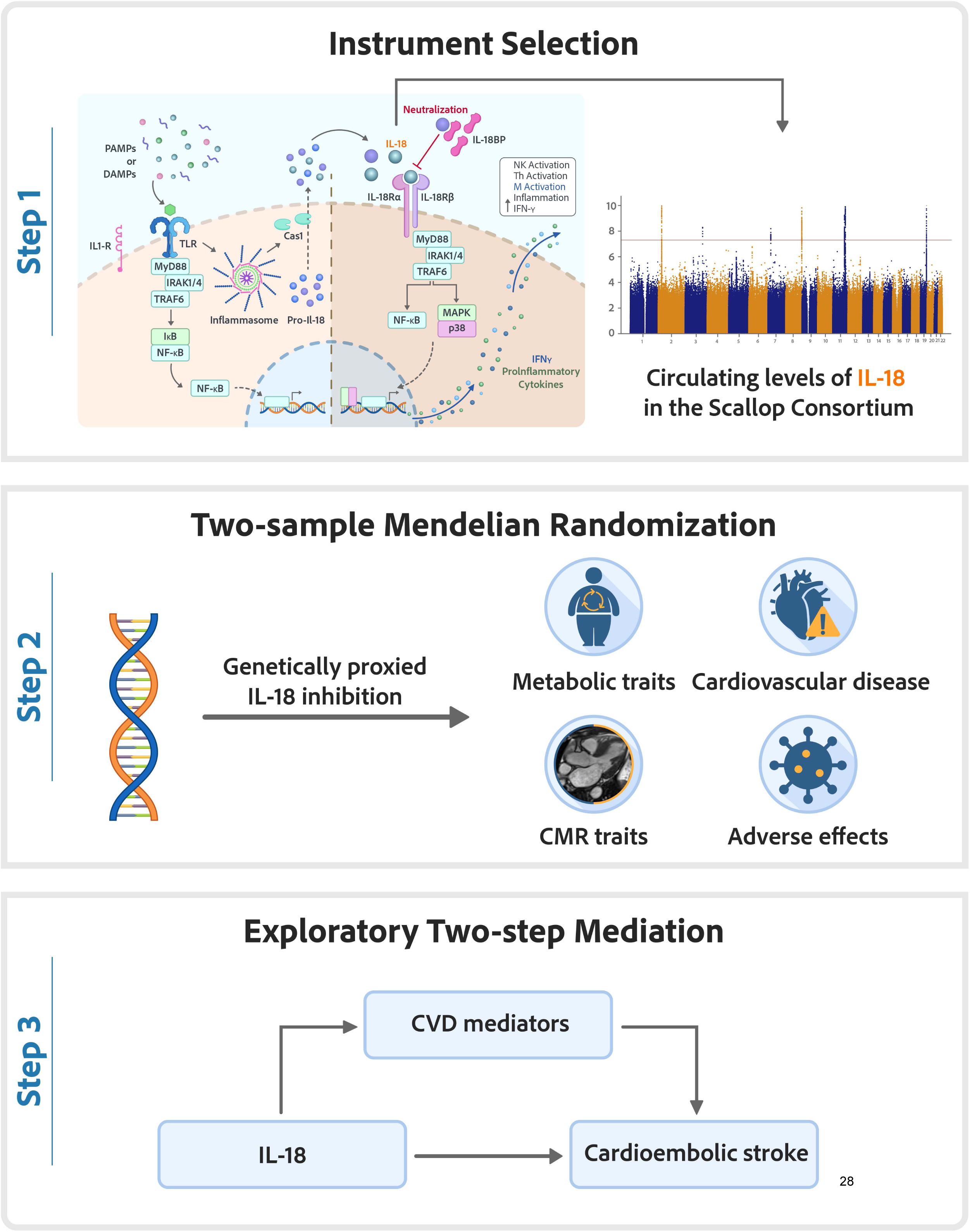
Overview of the study design. Stage 1 includes a graphical depiction of the interleukin-18 signaling pathway and a Manhattan plot illustrating the genome-wide association study of circulating concentration of interleukin-18 in the Scallop consortium. The red line indicates the genome-wide significance threshold (5 × 10⁻⁸).

Initially, we examined the validity of our genetic instrument as a pharmacological proxy for downregulated IL-18 signaling by assessing its impact on established downstream biomarkers: IL-6, C-reactive protein (CRP), interferon-gamma (IFN-γ), and tumor necrosis factor. These biomarkers were identified from pre-clinical^30^ and early-phase clinical studies of therapies targeting the IL-18 signaling pathway.^31,32^ Circulating levels of IL-1β were selected as a negative control because as IL-1β functions through an alternative signaling pathway, its levels are not expected to be affected by IL-18 signaling. Two-sample *cis-*MR was then used to elucidate the relationship between on-target reduced IL-18 signaling and the risk of cardiovascular disease. Subsequent analyses assessed the bidirectional effects between liability to diseases identified in primary analyses and circulating IL-18 levels. We undertook a mediation analysis to identify pathways that contribute to the effect of genetically predicted IL-18 blockade on cardioembolic stroke. Finally, we explored the effect of downregulated IL-18 signaling on longevity and a selection of clinically relevant outcomes to identify potential adverse outcomes associated with an impaired host immune response. We used parental age at death, a partially heritable trait, as an indirect measure of longevity.^33^ IL-18 downregulation may have off-target effects beyond cardiovascular outcomes, which is important for assessing the overall safety of therapies targeting IL-18. Given that IL-18 plays a role in immune surveillance, its inhibition could impair the body’s ability to detect and eliminate cancer cells, potentially increasing cancer risk. Existing MR evidence has already found an association between higher genetically predicted circulating IL-18 levels and an increased risk of lung cancer.^34^

#### Instrumental variable selection

Summary statistics for circulating IL-18 concentration were obtained from the Scallop Consortium’s genome-wide association meta-analysis of 90 Olink-measured proteins from 13 contributing cohorts containing 21,758 European individuals.^35^ We identified common (minor allele frequency >0.01) *cis-*genetic variants associated with circulating IL-18 concentration at genome-wide significance from within a 100kb flanking region of the *IL-18* gene (hg19: chromosome 11: 112013974-112034840). Variants were then clumped using a linkage disequilibrium (LD) threshold r^2^ <0.1 and a distance threshold of 100kb, utilizing data from the 1000 Genomes Project European reference panel. This methodology selected five partially correlated *cis-*variants with the strongest association with circulating IL-18 concentration within each LD region from around the *IL-18* gene, as described in Supplementary Table 2. Instrument strength was assessed through the calculation of an *F*-statistic and a mean *F*-statistic >10 was considered strong. Strong instruments provide sufficient statistical power to accurately test hypotheses. Weak instruments, which poorly predict the exposure, can lead to biased results, particularly in two-sample settings where they skew findings toward the null, reduce precision, and amplify bias due to violations of core instrumental variable assumptions.^29^ For bidirectional MR analyses, we created instruments for each cardiometabolic disease by selecting genetic variants associated with each outcome at genome-wide significance from throughout the genome. We clumped these variants using an LD threshold of r² < 0.001 and a distance threshold of 10,000 kb. We relaxed our significance threshold for HF to 5 × 10⁻⁷ due to a scarcity of genome-wide significant variants.

### Data Sources

Detailed information on all data sources used in this study can be found in Table 1, Supplementary Table 1, and the Supplementary Methods.

**Table 1.**
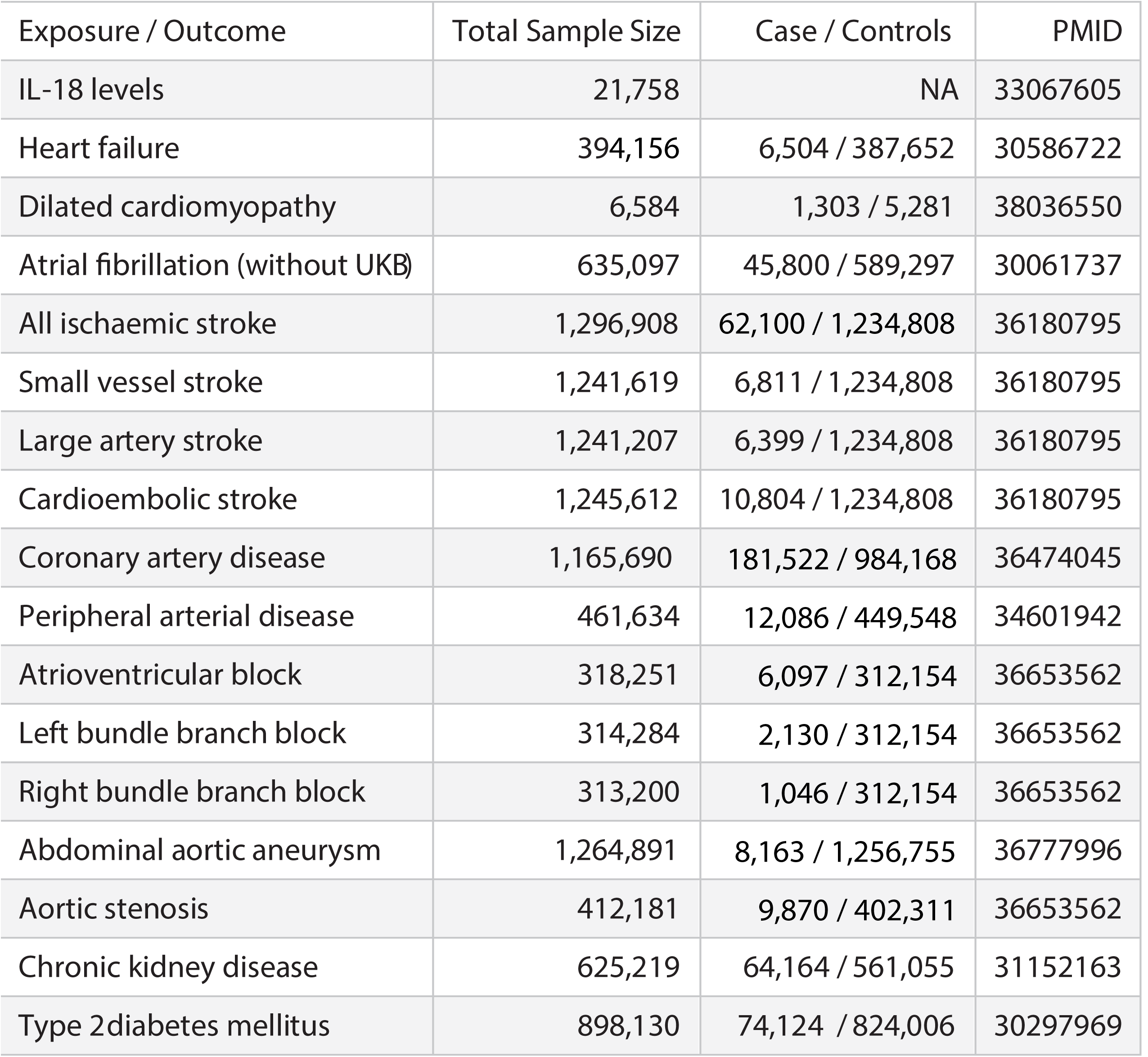
Overview of the cardiometabolic disease summary statistics used in this study.

| Exposure / Outcome | Total Sample Size | Case / Controls | PMID |
| --- | --- | --- | --- |
| IL-18 levels | 21,758 | NA | 33067605 |
| Heart failure | 394,156 | 6,504 / 387,652 | 30586722 |
| Dilated cardiomyopathy | 6,584 | 1,303 / 5,281 | 38036550 |
| Atrial fibrillation (without UKB) | 635,097 | 45,800 / 589,297 | 30061737 |
| All ischaemic stroke | 1,296,908 | 62,100 / 1,234,808 | 36180795 |
| Small vessel stroke | 1,241,619 | 6,811 / 1,234,808 | 36180795 |
| Large artery stroke | 1,241,207 | 6,399 / 1,234,808 | 36180795 |
| Cardioembolic stroke | 1,245,612 | 10,804 / 1,234,808 | 36180795 |
| Coronary artery disease | 1,165,690 | 181,522 / 984,168 | 36474045 |
| Peripheral arterial disease | 461,634 | 12,086 / 449,548 | 34601942 |
| Atrioventricular block | 318,251 | 6,097 / 312,154 | 36653562 |
| Left bundle branch block | 314,284 | 2,130 / 312,154 | 36653562 |
| Right bundle branch block | 313,200 | 1,046 / 312,154 | 36653562 |
| Abdominal aortic aneurysm | 1,264,891 | 8,163 / 1,256,755 | 36777996 |
| Aortic stenosis | 412,181 | 9,870 / 402,311 | 36653562 |
| Chronic kidney disease | 625,219 | 64,164 / 561,055 | 31152163 |
| Type 2diabetes mellitus | 898,130 | 74,124 / 824,006 | 30297969 |

### Statistical analysis

#### Mendelian randomization analyses

Causal estimates from MR analyses rely on three key assumptions: (i) relevance, where the genetic instrument must be associated with the exposure; (ii) independence, where there is no confounding of the relationship between the instruments and the outcome; and (iii) exclusion restriction, where the instrument affects the outcome solely through its effect on the exposure.^36^ Random-effects inverse-variance weighted (IVW) models were used in primary MR analyses. For all significant associations, we conducted a range of sensitivity analyses to assess for possible violations of the underlying assumptions of MR, including weighted median and mode to assess for consistency of results, MR Egger to assess for directional pleiotropy and MR PRESSO to adjust for the effect of potential outliers.^36^ We repeated all analyses using generalized inverse variance weighted MR, which adjusts for residual pairwise LD among variants included in our IL-18 instrument.^36^

#### Mediation analysis

We used two-step *cis-*MR to estimate the effect of the exposure on the outcome, conditional on the mediator (referred to as the “direct’ effect, reported as an adjusted OR with respective 95% CI). We then compared this adjusted effect to the unadjusted “total” effect to qualitatively assess whether substantial attenuation of the effect estimates occurs after conditioning on the mediator, which would suggest the presence of a mediating pathway. We only included traits with a biologically plausible mediation mechanism, and for which a valid genetic instrument could be constructed as a proxy for the potential mediator. We used the weighted median estimator for the mediation analysis if heterogeneity was detected in the inverse-variance weighted MR estimate. Two-step cis-MR is particularly useful for attenuating bias in situations where there are insufficient independent causal variants for methods like multivariable MR. Otherwise, it is analogous to conventional two-step MR mediation analysis.

As a secondary complementary approach, we obtained the “indirect” effect and proportion mediated through the product of coefficients method. The mediated proportion was obtained by dividing the indirect effect by the total effect and the SE for the indirect effect was calculated using the delta method.^37^ Given the binary nature of some mediators and outcomes in this study, caution is needed when interpreting the estimated indirect effect and proportion mediated. These estimates rely on methods that assume additive relationships among the exposure, mediator, and outcome, which may not be appropriate for binary outcomes.^38^

#### Colocalization and LD check

Bayesian colocalization was used to determine if levels of circulating IL-18 and our primary outcomes share a single causal variant at the IL-18 gene locus, which may indicate a causal pathway between the exposure and the outcome.^39^ Colocalization involves calculating posterior probabilities (PP) for five hypotheses to assess the likelihood that the genetic variants influencing two traits at a specific locus are overlapping or distinct. H0 suggests that there is no genetic association with either trait; H1 and H2 suggest that only the first or second trait has a genetic association; H3 suggests that both traits are associated but have different causal variants; and H4 suggests that both traits are associated and share a single causal variant. Colocalization analyses rely on a well-powered outcome GWAS and sufficient variant coverage in the region of interest.^39,40^ A posterior probability (PP) for H3 (distinct causal variant) and H4 (shared causal variant) less than 0.8, in the context of a statistically significant MR association, may signal an underpowered outcome GWAS to determine if the association is due to a shared causal variant or from a confounding variant in LD (i.e. horizontal pleiotropy).

LD Check is an alternative method for co-localization when variant coverage is insufficient for Bayesian colocalization.^40^ LD Check estimates the LD between the sentinel IL-18 variant and the top 30 variants from the same genomic region in each outcome study. Causality with the sentinel IL-18 variant is suggested if at least one of the top 30 SNPs exhibits strong LD (r^2^ >0.8).^40^ This indicates that these SNPs are highly correlated and may tag the same causal variant. Such findings provide further evidence of a shared genetic basis, complementing traditional colocalization methods and the MR findings. While traditional colocalization methods estimate the probability of a shared causal variant through statistical modeling, LD checks offer a more direct assessment by leveraging LD structure. This approach is particularly valuable when the statistical power of colocalization or other methods is limited due to low SNP coverage."

#### Reporting and packages

Results from MR analyses are reported as odds ratios (OR) for binary outcomes and standard deviation (SD) difference for continuous outcomes per SD lower in genetically predicted IL-18 levels. Results from bidirectional MR analyses are reported as the effect of a one-unit increase in the log odds of genetic liability for each cardiometabolic disease. We applied the Benjamini-Hochberg method to control for the false discovery rate (FDR) across all phenotypes and report the unadjusted and FDR-adjusted p-values. Associations with a nominal p-value <0.05 and a Benjamini-Hochberg adjusted p-value >0.05 were deemed suggestive. Associations with a Benjamini-Hochberg adjusted p-value <0.05 were considered significant. Analyses were performed in R (v4.2.1; R Foundation for Statistical Computing) with the TwoSampleMR, MendelianRandomisation, Coloc and TwoStepCisMR packages.

### Standard Protocol Approvals, Registrations, and Patient Consents

All contributing studies obtained appropriate informed consent from participants and received ethical approval before commencement. This study was conducted in adherence to the guidelines for Strengthening the Reporting of Observational Studies in Epidemiology – Mendelian randomization (STROBE-MR), as detailed in the Supplement.

## Data availability

This study utilizes publicly available, summary-level GWAS data that can be accessed through the GWAS Catalogue (https://www.ebi.ac.uk/gwas/home) and the Integrative Epidemiology Unit (IEU) OpenGWAS project (https://gwas.mrcieu.ac.uk/) or specific cohort portals, including The Scallop Consortium (https://zenodo.org/records/2615265), The Centre for Statistical Genetics (https://csg.sph.umich.edu/willer/public/afib2018/), Pan-UK Biobank (https://pan.ukbb.broadinstitute.org/), The Human Genetics Amplifier (HuGeAMP) (https://kp4cd.org/node/169), The Usher Institute of Population Health Sciences and Informatics (https://datashare.ed.ac.uk/handle/10283/3209), The Cardiovascular Disease Knowledge Portal (https://cvd.hugeamp.org/downloads.html), The NBDC Human Database (https://humandbs.dbcls.jp/en/), The Complex Trait Genetics Lab (https://cncr.nl/research/summary_statistics/) and FinnGen (https://www.finngen.fi/en/access_results).

## Results

### Genetically downregulated IL-18 signaling and cardiometabolic biomarkers

Our instrument for lower genetically downregulated IL-18 signaling, composed of 5 variants, had a mean *F*-statistic of 124, indicating an appropriate average instrument strength, and explained 3.64% of the total variation of circulating IL-18 levels in the Scallop Consortium. Lower genetically predicted IL-18 levels were associated with several downstream biomarkers of IL-18 signaling, suggesting that our instruments represented a proxy for IL-18 signaling downregulation. The downstream biomarkers that were significantly associated with genetically proxied IL-18 levels included C-reactive protein (CRP) (SD change -0.02, 95% CI -0.03, -0.02, p_fdr_ <0.001), tumor necrosis factor (SD change -0.19, 95% CI -0.25, -0.14, p_fdr_ <0.001), interferon-gamma (IFN-y) (SD change -0.15, 95% CI -0.22, -0.08, p_fdr_ <0.001) and CXCL10 (SD change -0.13, 95% CI -0.16, -0.09, p_fdr_ <0.001), but not IL-1β or IL-6 (Figure 2). Lower genetically predicted IL-18 levels were associated with lower fasting insulin (SD change -0.02, 95% CI -0.04, -0.01, p_fdr_ = 0.023), higher diastolic blood pressure (SD change 0.02, 95% CI 0.02, 0.03, p_fdr_ =<0.001) and body mass index (SD change 0.02, 95% CI 0.02, 0.04, p_fdr_ =0.015), but had no association with lipid subfractions or fasting glucose.

**Figure 2.**
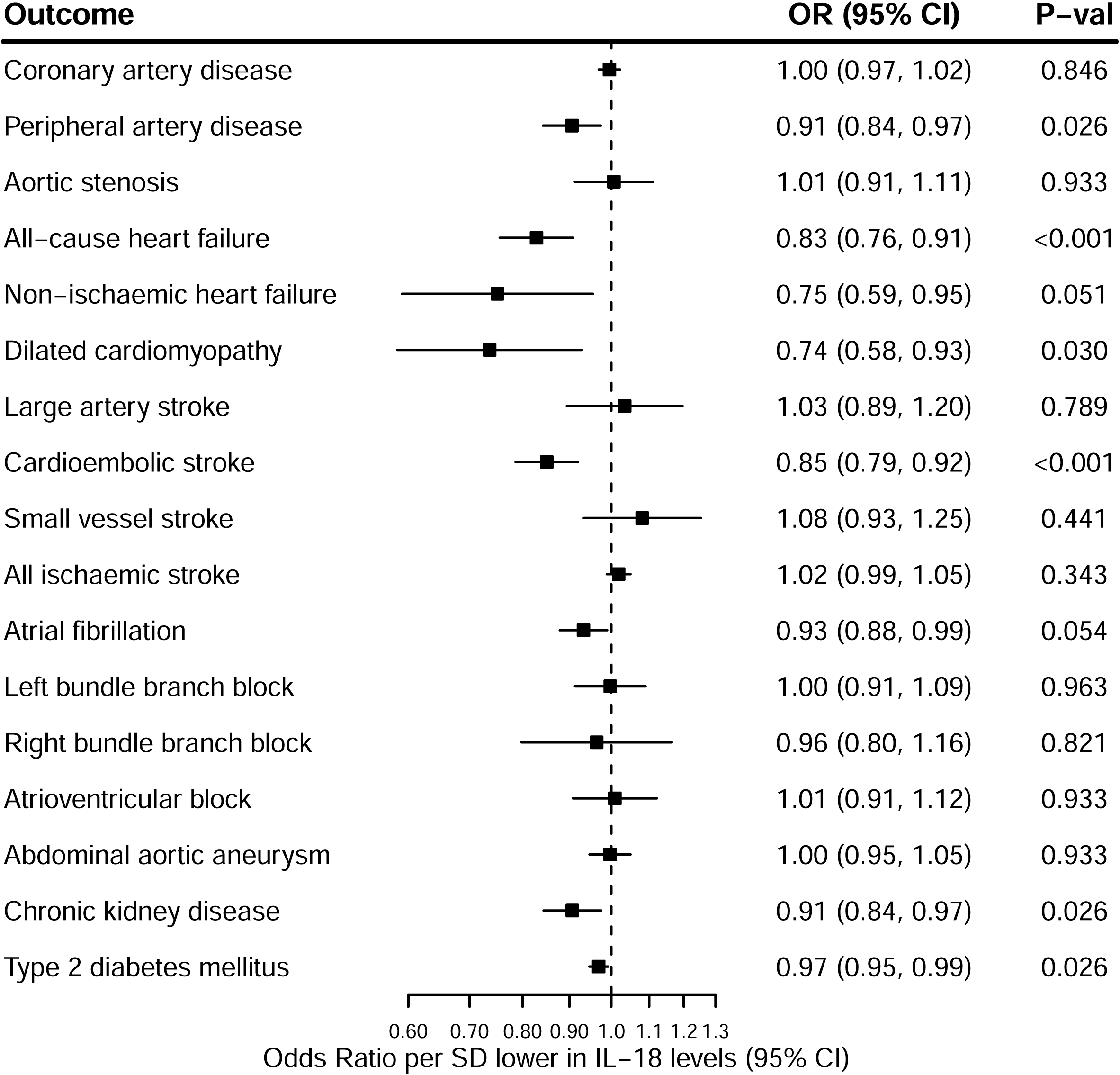
The association between genetically downregulated IL-18 signaling and cardiometabolic biomarkers. Shown are the results derived from random-effects inverse-variance weighted Mendelian randomization analyses. OR indicates odds ratio; CI, confidence interval; SD, standard deviation; FDR, false discovery rate; CXCL10, C-X-C motif chemokine ligand 10.

### Genetically downregulated IL-18 signaling and cardiometabolic diseases

Lower genetically predicted IL-18 levels (1 SD decrease in IL-18 levels) were associated with 15% lower odds of cardioembolic stroke (OR 0.85, 95% CI 0.79-0.92, p_fdr_ <0.001, Figure 3) and 9% lower odds of peripheral arterial disease (OR 0.91, 95% CI 0.84-0.97, p_fdr_ =0.026), but had no association with other stroke subtypes, coronary artery disease or aortic stenosis. In UKB, lower genetically predicted IL-18 levels were also associated with reduced odds of both all-cause HF (OR 0.83, 95% CI 0.75-0.91, p_fdr_ <0.001) and dilated cardiomyopathy (OR 0.74, 95% CI 0.58-0.93, p_fdr_ =0.030). When assessing related cardio-metabolic diseases, lower genetically predicted IL-18 levels were associated with lower odds of chronic kidney disease (CKD) (OR 0.91, 95% CI 0.84-0.99, p_fdr_=0.026), and type 2 diabetes mellitus (T2DM) (OR 0.97, 95% CI 0.95-0.99, p_fdr_ = 0.026). Additionally, there was a trend towards an association between lower genetically predicted IL-18 levels and reduced risk of AF (OR 0.94, 95% CI 0.89-0.99, p_fdr_ = 0.054), which did not maintain significance following false discovery rate adjustment. Associations for HF were successfully replicated in a non-overlapping sample of American cohort studies (OR 0.85, 95% CI 0.75-0.95, p =0.004, Supplementary Table 3). For AF, results were directionally similar but did not reach statistical significance in the UK Biobank (UKB) (OR 0.96, 95% CI 0.92-1.00, p = 0.061).

**Figure 3.**
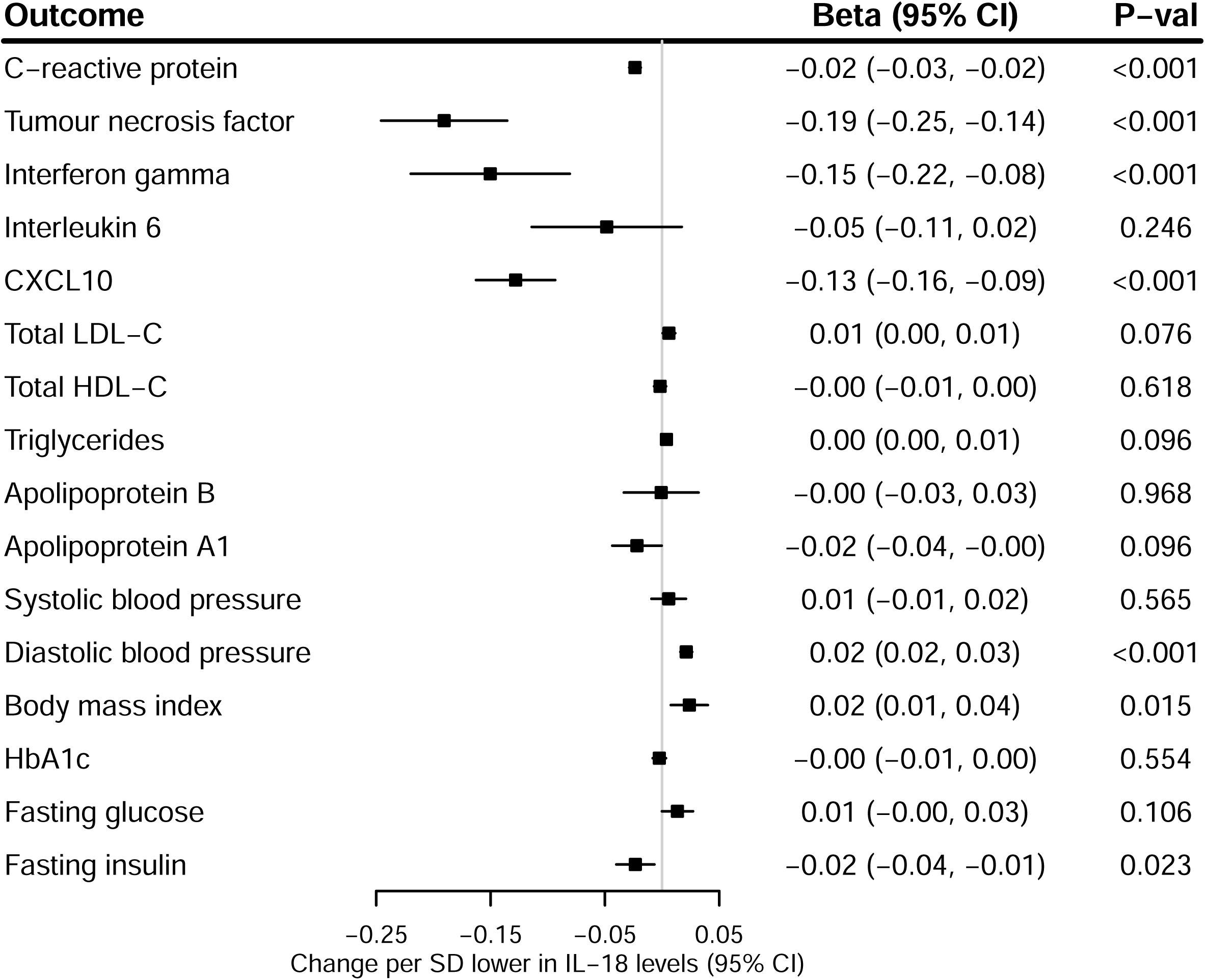
The association between genetically downregulated IL-18 signaling and cardiometabolic diseases. Shown are the results derived from random-effects inverse-variance weighted Mendelian randomization analyses. OR indicates odds ratio; CI, confidence interval; SD, standard deviation.

In colocalization analyses between IL-18 and AF, peripheral arterial disease, HF, cardioembolic stroke, CKD and T2DM, the posterior probabilities of *H3* and *H4* were less than 0.8 and this along with the low minimum p-values in the *IL-18* gene region in each of the outcome summary statistics implies suboptimal power for colocalization, the presence of confounding by LD or multiple causal variants within the gene region. We subsequently conducted an LD Check analysis and obtained the LD correlation between the sentinel variant (rs5744249) from the *IL-18* gene locus and the top 30 variants within 100kb of rs5744249 with the lowest p-values from the cardioembolic stroke, AF, peripheral arterial disease, HF, CKD and T2DM summary statistics. At least one top *cis-*variant from each outcome dataset, except for peripheral arterial disease, exhibited strong LD (r^2^ > 0.8) with the sentinel variant. This suggests potential causal associations between IL-18 and cardioembolic stroke, AF, HF, CKD and T2DM.

### Genetically downregulated IL-18 signaling and cardiac magnetic resonance imaging traits

In UKB, there were directionally consistent improvements in all measures of cardiac structure and function with lower genetically predicted IL-18 levels as displayed in Figure 3, with the exception of left and right ventricular ejection fractions. Following adjustment for multiple testing, lower genetically predicted IL-18 levels were associated with a reduction in left atrial size (SD change -0.02, 95% CI -0.03, -0.00, p_fdr_ = 0.023) and left ventricular end-systolic volume (SD change -0.08, 95% CI -0.13, -0.02, p_fdr_ = 0.038).

### Genetically downregulated IL-18 signaling and potential adverse effects

Lower genetically predicted IL-18 levels were associated with a longer parental lifespan, an indirect measure of longevity (SD change 0.02 years, 95% CI 0.01-0.04, p_fdr_ = 0.005), and reduced risk of a range of autoimmune diseases, including atopic dermatitis (OR 0.86, 95% CI 0.83-0.90, p_fdr_ <0.001), psoriasis (OR 0.84-0.89, p_fdr_ <0.001), asthma (OR 0.90, 95% CI 0.86-0.94, p_fdr_<0.001), and rheumatoid arthritis (OR 0.85, 95% CI 0.81-0.90, p_fdr_ < 0.001, Figure 4). There was also a trend toward an association between lower genetically predicted IL-18 levels and reduced risk of ulcerative colitis (OR 0.88, 95% CI 0.79-0.98, p_fdr_ = 0.051), which did not maintain significance following false discovery rate adjustment. While lower genetically predicted IL-18 levels were not significantly associated with an increased risk of infection, it was associated with higher odds of lung cancer (OR 1.15, 95% CI 1.11-1.20, p_fdr_ <0.001) and multiple sclerosis (OR 1.17, 95% CI 1.03-1.32, p_fdr_ = 0.039).

**Figure 4.**
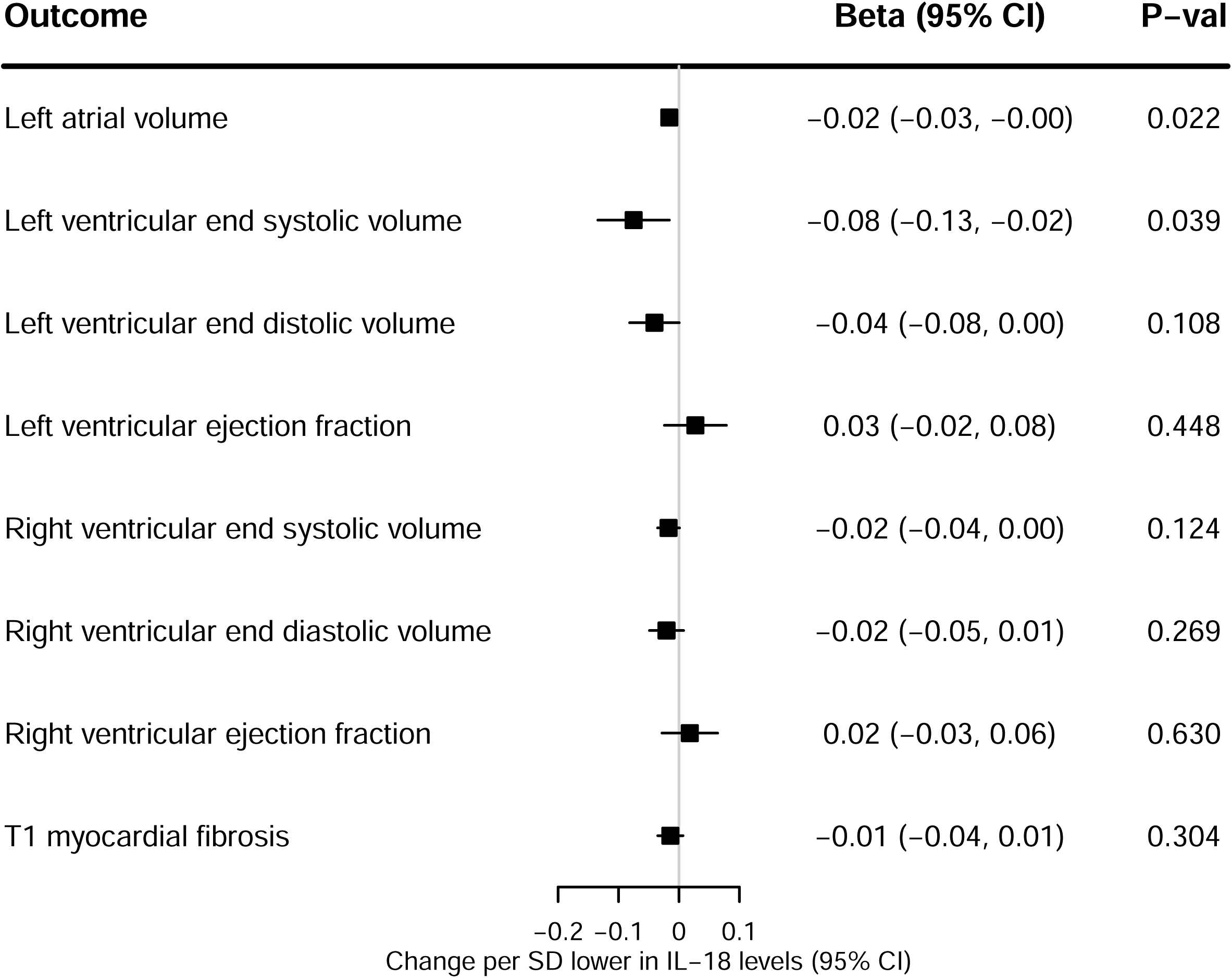
The association between genetically downregulated IL-18 signaling and cardiac magnetic resonance imaging traits. Shown are the results derived from random-effects inverse-variance weighted Mendelian randomization analyses. OR indicates odds ratio; CI, confidence interval; SD, standard deviation.

**Figure 5.**
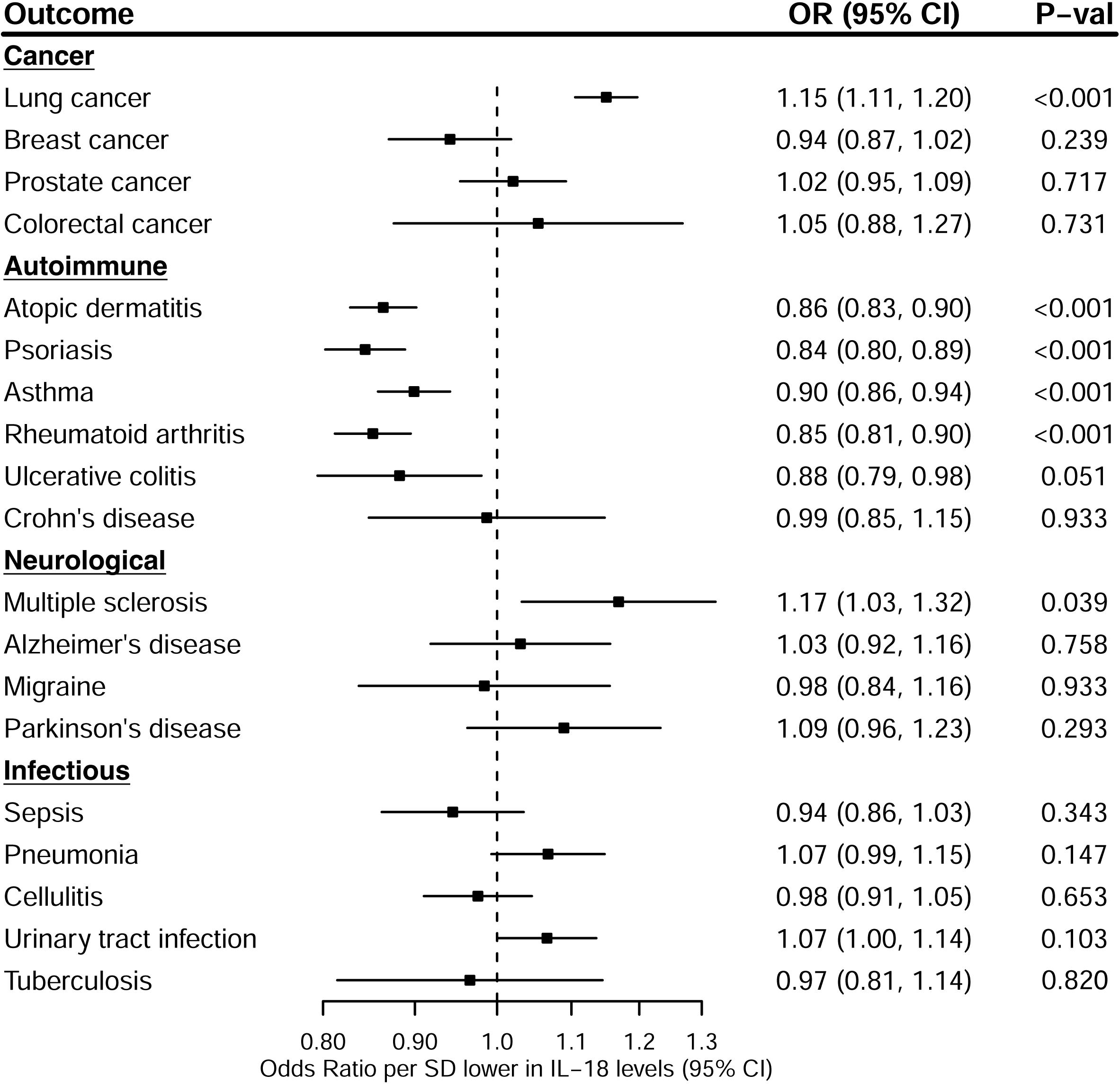
The association between genetically downregulated IL-18 signaling and oncological, autoimmune, neurological, and infectious diseases. Shown are the results derived from random-effects inverse-variance weighted Mendelian randomization analyses. OR indicates odds ratio; CI, confidence interval; SD, standard deviation.

### Bidirectional relationship between cardiometabolic diseases and IL-18 levels

In reverse MR analyses, genetic liability to cardioembolic stroke was not significantly associated with circulating levels of IL-18 (β representing the effect of one unit increase in the log odds of genetic liability for cardioembolic stroke -0.03, 95% CI -0.08-0.02, p = 0.267). Similarly, circulating levels of IL-18 were not affected by genetic liability to AF (β 0.00, 95% CI -0.03-0.04, p 0.879), HF (β -0.03, 95% CI -0.22-0.16, p =0.743), CKD (β 0.01, 95% CI -0.07-0.06, p = 0.821), peripheral arterial disease (β 0.07, 96% CI -0.26-0.39, p = 0.686), or higher BMI (β 0.05, 95% CI -0.01-0.12, p = 0.104).

### Mediation analyses

We performed separate mediation analyses to quantify the extent to which each downstream cardiometabolic trait mediates the relationship between IL-18 signaling and cardioembolic stroke. The association between IL-18 and cardioembolic stroke (unadjusted OR 0.86, 95% CI 0.79-0.92, p < 0.001) showed attenuation after adjustment for genetic liability to atrial fibrillation (adjusted OR 0.89, 95% CI 0.80-0.98, p = 0.018), suggesting that a significant proportion of the causal effect of IL-18 on cardioembolic stroke is mediated through atrial fibrillation. C-reactive protein and genetic liabilities to HF, CKD, T2DM and obesity had minimal impact (Supplementary Table 5.1-5.2).

### Sensitivity analyses

The MR estimates from generalized IVW, weighted median, weighted mode, MR Egger, and MR PRESSO are reported in Supplementary Tables 6.1-6.24. No heterogeneity was observed across all MR analyses, as indicated by Cochrane’s Q and Rucker’s Q statistics. Estimates were consistent with primary results when using generalized IVW MR to model residual LD, and directionally similar across weighted median and weighted mode methods. The p-values for the MR Egger intercept test and the global MR PRESSO test were not significant, suggesting an absence of marked pleiotropy.

## Discussion

Leveraging data from large-scale genomic, proteomic and imaging studies, we have demonstrated new and important evidence linking IL-18 signaling to HF, peripheral arterial disease, atrial cardiomyopathy, and cardioembolic stroke. Lower genetically predicted IL-18 levels were associated with reduced concentrations of several downstream inflammatory cytokines and exhibited protective effects on cardiac structure and function. Genetic liability to AF and HF mediated a significant proportion of the risk-lowering effect of IL-18 signaling on cardioembolic stroke. Lower genetically predicted IL-18 levels were associated with a favorable metabolic profile overall, with a lower risk of T2DM, CKD, autoimmune disease, and enhanced longevity.

The present data are supported by several lines of evidence from experimental models, observational studies, and emerging data from RCTs implicating the NLRP3-inflammasome and IL-18 activation in HF and AF. The NLRP3-inflammasome is activated by pro-inflammatory mediators, such as IL-1β and TNF-alpha, damage-associated molecular patterns (DAMPs) secondary to ischemia-reperfusion injury, and angiotensin II over-expression from renin-angiotensin activation.^4,21,41^ IL-18 activation ensues secondary to NLRP3 inflammasome formation in leukocytes, endothelial cells, fibroblasts, and cardiomyocytes.^21^ The binding of IL-18 to its receptor triggers the recruitment of tumor necrosis factor receptor-associated factor-6 (TRAF6), and the activation of nuclear factor Kappa B (NF-κB) and p38 mitogen-activated protein kinase (MAPK).^21^ This cascade results in the synthesis of secondary inflammatory mediators, including IFN-γ and TNF-α.^30^ In-vitro and animal models have demonstrated that disrupting the IL-18 signaling pathway can reduce myocardial hypertrophy, improve contractile function, and decrease fibrosis.^21,22^ IL-18 mediates β-adrenergic receptor stimulation-induced macrophage infiltration, cardiac inflammation, and fibrosis.^20^ IL-18 also triggers apoptosis in microvascular endothelial cells through activation of NF-κB and upregulates the expression of osteopontin, fibronectin, and matrix metalloproteinase 2 (MMP-2) in human cardiac fibroblasts.^21^ In humans, IL-18 levels increase in proportion to disease progression in both HF^23^ and AF.^42,43^ In the Atherosclerosis Risk in Communities (ARIC) study circulating IL-18 in mid-life was associated with future risk of HF.^24^ Similarly, Henry et al.’s meta-analysis of four population-based studies demonstrated an association between IL-18 levels and risk of incident HF.^27^

Our results suggest that the IL-18 signaling pathway is a viable treatment target in AF, HF and cardioembolic stroke. Our data were derived from non-overlapping cohorts and demonstrated biologically consistent results, implicating IL-18 at the center of multiple downstream pro-inflammatory pathways, cardiac remodeling, AF, HF, and ultimately cardioembolic stroke. Our two-step *cis-*MR analysis suggests that the reduced risk of cardioembolic stroke associated with downregulated IL-18 signaling is likely mediated through its protective effects against lifetime cardiac remodeling and AF. Our findings are supported by Schmidt et al.’s recent cis-MR study, which showed that targeting the IL-18 receptor is associated with four left ventricular CMR traits and risk of dilated cardiomyopathy in UKB.^44^ Additionally, van Vugt et al. replicated an association between IL-18R and AF across the deCODE, AGES-Reykjavik, and INTERVAL cohorts.^45^ In their candidate gene study, Wang et al. demonstrated that three variants associated with IL18 gene expression increased risk of AF and left atrial enlargement.^46^

The global burden of stroke, HF and AF is increasing.^47,48^ There is a pressing need to identify and develop disease-modifying therapies that prevent stroke and slow disease progression in both HF and atrial cardiopathy. In atherosclerosis, colchicine and canakuinumab have both shown benefit in reducing major adverse cardiovascular events (MACE) in coronary artery disease.^1,2^ Although colchicine did not reduce the risk of MACE after non-cardioembolic stroke, a 20% reduced risk was observed in an on-treatment analysis suggesting a possible biological effect.^3^ Apart from the treatment of atherosclerosis, there is also increasing interest in targeting inflammation in HF and AF. Whilst there is some evidence that colchicine reduces the risk of postoperative AF following cardiac surgery and decreases AF recurrence after pulmonary vein isolation or ablation^49^, a recent RCT was neutral.^12^ Currently, no licensed anti-inflammatory therapies have been proven to prevent hard outcomes in HF. RCTs investigating the potential of TNFα inhibition were neutral.^10,11^ Anakinra, an anti-IL-1 monoclonal antibody, improved several surrogate markers of cardiac function but failed to significantly reduce hospitalization for HF or death.^50^ While genetic studies have identified the IL-6 signaling pathway as a potential druggable target for AF, they do not support a causal role in HF or cardioembolic stroke.^16^ Consequently, future experimental work and RCTs should now focus on the potential benefit of IL-18 pathway suppression in the treatment of HF and the prevention of cardioembolic stroke.

Our results suggest that IL-18 inhibition would likely have a favourable cardio-metabolic profile. Downregulated IL-18 signaling reduced risk of CKD and T2DM, which are key risk factors for cardiovascular disease. However, IL-18 suppression was associated with marginal increases in diastolic blood pressure and BMI, with minimal effects on lipid fractions and systolic blood pressure. The clinical significance of these findings will need to be further explored in future studies. The MR estimate for the effect of lower IL-18 signaling on LDL cholesterol was small and did not reach significance after adjusting for multiple testing (SD change 0.01, 95% CI 0.00-0.01, p = 0.08). This relationship may not be biologically relevant and could also be influenced by the large size of the outcome summary statistics. A similar phenomenon may explain the findings for diastolic blood pressure and BMI. However, pharmacological therapies can exhibit pleiotropic effects that appear paradoxical despite their beneficial effect on certain conditions. For example, statins reduce the risk of atherosclerotic cardiovascular disease by lowering LDL cholesterol levels, while also increasing pro-atherogenic Lp(a) levels.^51^ Similarly, while pre-clinical and MR studies suggest that inhibiting IL-6 signaling is protective against CVD, clinical trials have shown that IL-6 receptor blockers, such as tocilizumab, increase total cholesterol and LDL cholesterol levels, which would traditionally be expected to raise CVD risk.^52^

We show that downregulated IL-18 signaling is associated with lower risk of atopic disease, which supports the rationale for ongoing clinical trials of IL-18 signaling-blocking therapies in atopic dermatitis. Promising agents for potential repurposing in cardiovascular disease include GSK1070806 and Camoteskimab, which are human monoclonal antibodies targeting IL-18, as well as tadekinig α, a recombinant human IL-18 binding protein that sequesters IL-18. Immunomodulatory therapies carry potential risks, notably with respect to host response to infections and malignancy. Our study shows that downregulated IL-18 signaling increases the risk of lung cancer, but not other cancers or infection and is associated with enhanced lifespan, suggesting a net positive effect on all-cause mortality. The relationship between IL-18 and lung cancer has been identified in previous MR studies,^34^ and a phase 2 clinical trial is assessing the effect of a decoy-resistant variant of recombinant IL-18 in a range of solid tumours including non-small cell lung cancer.^53^ IL-18 plays a crucial role in cellular immunity by activating T-helper cells, which stimulate an effective antitumor immune response. Our results suggest potential cardiometabolic risks associated with using recombinant IL-18 therapies for cancer treatment, highlighting safety outcomes that should be monitored in clinical trials. Importantly, we show that reduced IL-18 signaling is associated with enhanced lifespan, indicative of a net positive effect on all-cause mortality. Our study suggests a potential causal link between IL-18 inhibition and risks of lung cancer and multiple sclerosis. However, given the relative rarity of these conditions compared to the high global burden of stroke and cardiomyopathy, the overall benefits of IL-18 inhibition, particularly in reducing cardiovascular risk, may outweigh potential risks. Personalized treatment strategies and risk stratification could help mitigate adverse effects, particularly in individuals with risk factors for lung cancer or multiple sclerosis. Further clinical research is necessary to confirm these findings and ensure the long-term safety of IL-18 inhibitors in diverse populations

Previous data from experimental models and some epidemiological studies suggested that IL-18 was associated with atherosclerosis and coronary disease.^26,54^ For these reasons, we expected to see an association with atherosclerotic endpoints. However, apart from a modest association with peripheral arterial disease that did not have LD Check support, we did not find any compelling evidence that IL-18 signaling is causally associated with atherosclerotic events. Previous observational data may have been the result of reverse causality whereby IL-18 expression may have been a consequence of atherosclerotic disease progression rather than a causal factor. Alternatively, it is also possible that our genetic instrument does not fully capture compensatory mechanisms that occur within vascular tissues, such as the upregulation of IL-18 binding protein, which inhibits IL-18 from binding to its receptors and modulates its biological activity. On the contrary, prior MR studies have implicated IL-6 signaling in coronary artery disease and atherosclerotic stroke.^16^ Together with data from CANTOS suggesting the potential benefit of IL-1β-IL-6 axis suppression,^2^ our analysis supports the rationale of ongoing trials focusing on the high potential of IL-6 pathway inhibition in atherosclerosis.^17^

Our study has several strengths. The MR study design minimizes bias from reverse causation and environmental confounders, while cis-MR provides an evaluation of the effects of directly perturbing a drug target on disease outcomes. We leveraged large multi-omic datasets to uncover new insights into how downregulated IL-18 signaling influences cardiovascular disease. Moreover, we developed a robust genetic cis-instrument that mirrors the effects of reduced IL-18 signaling on downstream biomarkers and conducted a comprehensive set of mediation and sensitivity analyses to improve confidence in our findings. We were also able to validate our findings for HF in independent cohorts.

We acknowledge some limitations. Although we utilized large genomic consortia for outcome summary statistics, our analyses may have been underpowered to detect associations with rarer phenotypes. Effect sizes derived from drug-target MR studies reflect lifelong perturbation of the drug target rather than the short-term treatment durations typically seen in clinical practice, potentially leading to significant differences in the observed benefits. Two-sample MR methods assume a linear relationship between genetically proxied exposures and outcomes. In cases of non-linearity, the MR estimates reflect a population average effect rather than capturing potentially varying effects across different levels of exposure. Similarly, in clinical practice, the biological response to drugs often requires surpassing a specific therapeutic dose, a detail that two-sample MR studies might not capture due to their reliance on average effects. Two-step *cis*-MR relies on several assumptions including linearity, and may yield unreliable estimates when the mediator or outcome is not continuous, potentially impacting the accuracy of our effect size estimates and the interpretation of IL-18 signaling mediation on cardioembolic stroke through AF, HF, CKD, and T2DM. Additionally, using genetic liability as a proxy for disease may complicate interpretation, as liability does not equate to actual diagnosis. Our analyses were limited to individuals of European ancestry to minimize confounding by population stratification and the potential impact of downregulated IL-18 signaling on risk of cardio-vascular disease in non-European ancestries remains to be explored. Finally, our MR estimates reflect the impact of downregulated IL-18 signaling on disease development. However, it remains unclear how appliable these findings are to disease progression in individuals with established disease, who are more likely to be prioritized for treatment and included in clinical trials.

In conclusion, this study used large-scale multi-omic data to explore the effects of genetically downregulated IL-18 signaling on cardiovascular disease. We provide compelling data which suggests the IL-18 pathway is causally implicated in the evolution of cardiomyopathy, from structural remodelling through to an increased risk of HF and cardioembolic stroke. These data strongly suggest that IL-18 represents a viable target for the prevention and treatment of stroke and heart failure, with a favorable cardiometabolic and autoimmune profile, that warrants further exploration in clinical trials.

## Abbreviations

HF: Heart failure
NLRP: NOD-like receptor family pyrin domain-containing
AF: Atrial fibrillation
IL-18: Interleukin-18
CMR: Cardiac magnetic resonance
CRP: C-reactive protein
IFN-γ: Interferon-gamma
CVD: cardiovascular disease
MR: Mendelian randomization
GWAS: Genome-wide association study

## References

1. Xia M, Yang X, Qian C. Meta-analysis Evaluating the Utility of Colchicine in Secondary Prevention of Coronary Artery Disease. Am J Cardiol. 2021;140:33–38. doi: 10.1016/j.amjcard.2020.10.043

2. Ridker PM, Everett BM, Thuren T, MacFadyen JG, Chang WH, Ballantyne C, Fonseca F, Nicolau J, Koenig W, Anker SD, et al. Antiinflammatory Therapy with Canakinumab for Atherosclerotic Disease. New England Journal of Medicine. 2017;377:1119–1131.

3. Kelly P, Lemmens R, Weimar C, Walsh C, Purroy F, Barber M, Collins R, Cronin S, Czlonkowska A, Desfontaines P. Long-term colchicine for the prevention of vascular recurrent events in non-cardioembolic stroke (CONVINCE): a randomised controlled trial. The Lancet. 2024.

4. Yao C, Veleva T, Scott L, Cao S, Li L, Chen G, Jeyabal P, Pan X, Alsina KM, Abu-Taha I, et al. Enhanced Cardiomyocyte NLRP3 Inflammasome Signaling Promotes Atrial Fibrillation. *Circulation (New York*, NY). 2018;138:2227–2242. doi: 10.1161/CIRCULATIONAHA.118.035202

5. McKechnie DGJ, Papacosta AO, Lennon LT, Welsh P, Whincup PH, Wannamethee SG. Inflammatory Markers and Incident Heart Failure in Older Men: the Role of Nt-Probnp. Biomarkers in Medicine. 2021;15:413–425. doi: 10.2217/bmm-2020-0669

6. Szabo TM, Frigy A, Nagy EE. Targeting Mediators of Inflammation in Heart Failure: A Short Synthesis of Experimental and Clinical Results. Int J Mol Sci. 2021;22. doi: 10.3390/ijms222313053

7. Conen D, Ridker PM, Everett BM, Tedrow UB, Rose L, Cook NR, Buring JE, Albert CM. A multimarker approach to assess the influence of inflammation on the incidence of atrial fibrillation in women. Eur Heart J. 2010;31:1730–1736. doi: 10.1093/eurheartj/ehq146

8. Aviles RJ, Martin DO, Apperson-Hansen C, Houghtaling PL, Rautaharju P, Kronmal RA, Tracy RP, Van Wagoner DR, Psaty BM, Lauer MS, et al. Inflammation as a risk factor for atrial fibrillation. Circulation. 2003;108:3006–3010. doi: 10.1161/01.CIR.0000103131.70301.4F

9. Aulin J, Siegbahn A, Hijazi Z, Ezekowitz MD, Andersson U, Connolly SJ, Huber K, Reilly PA, Wallentin L, Oldgren J. Interleukin-6 and C-reactive protein and risk for death and cardiovascular events in patients with atrial fibrillation. Am Heart J. 2015;170:1151–1160. doi: 10.1016/j.ahj.2015.09.018

10. Mann DL, McMurray JJ, Packer M, Swedberg K, Borer JS, Colucci WS, Djian J, Drexler H, Feldman A, Kober L, et al. Targeted anticytokine therapy in patients with chronic heart failure: results of the Randomized Etanercept Worldwide Evaluation (RENEWAL). Circulation. 2004;109:1594–1602. doi: 10.1161/01.Cir.0000124490.27666.B2

11. Chung ES, Packer M, Lo KH, Fasanmade AA, Willerson JT. Randomized, double-blind, placebo-controlled, pilot trial of infliximab, a chimeric monoclonal antibody to tumor necrosis factor-alpha, in patients with moderate-to-severe heart failure: results of the anti-TNF Therapy Against Congestive Heart Failure (ATTACH) trial. Circulation. 2003;107:3133–3140. doi: 10.1161/01.Cir.0000077913.60364.D2

12. Benz AP, Amit G, Connolly SJ, Singh J, Acosta-Vélez JG, Conen D, Deif B, Divakaramenon S, McIntyre WF, Mtwesi V, et al. Colchicine to Prevent Atrial Fibrillation Recurrence After Catheter Ablation: A Randomized, Placebo-Controlled Trial. Circulation: Arrhythmia and Electrophysiology. 2024;17:e01238. doi: doi:10.1161/CIRCEP.123.012387

13. Abbate A, Toldo S, Marchetti C, Kron J, Van Tassell BW, Dinarello CA. Interleukin-1 and the inflammasome as therapeutic targets in cardiovascular disease. Circulation research. 2020;126:1260–1280.

14. Kaptoge S, Seshasai SR, Gao P, Freitag DF, Butterworth AS, Borglykke A, Di Angelantonio E, Gudnason V, Rumley A, Lowe GD, et al. Inflammatory cytokines and risk of coronary heart disease: new prospective study and updated meta-analysis. Eur Heart J. 2014;35:578–589. doi: 10.1093/eurheartj/eht367

15. Papadopoulos A, Palaiopanos K, Bjorkbacka H, Peters A, de Lemos JA, Seshadri S, Dichgans M, Georgakis MK. Circulating Interleukin-6 Levels and Incident Ischemic Stroke: A Systematic Review and Meta-analysis of Prospective Studies. Neurology. 2021. doi: 10.1212/WNL.0000000000013274

16. Georgakis MK, Malik R, Gill D, Franceschini N, Sudlow CLM, Dichgans M, Invent Consortium CIWG. Interleukin-6 Signaling Effects on Ischemic Stroke and Other Cardiovascular Outcomes: A Mendelian Randomization Study. Circ Genom Precis Med. 2020;13:e002872. doi: 10.1161/CIRCGEN.119.002872

17. Ridker PM, Rane M. Interleukin-6 Signaling and Anti-Interleukin-6 Therapeutics in Cardiovascular Disease. Circ Res. 2021;128:1728–1746. doi: 10.1161/CIRCRESAHA.121.319077

18. McCabe JJ, Walsh C, Gorey S, Harris K, Hervella P, Iglesias-Rey R, Jern C, Li L, Miyamoto N, Montaner J, et al. C-Reactive Protein, Interleukin-6, and Vascular Recurrence After Stroke: An Individual Participant Data Meta-Analysis. Stroke. 2023. doi: 10.1161/STROKEAHA.122.040529

19. Toldo S, Mezzaroma E, O’Brien L, Marchetti C, Seropian IM, Voelkel NF, Van Tassell BW, Dinarello CA, Abbate A. Interleukin-18 mediates interleukin-1-induced cardiac dysfunction. Am J Physiol Heart Circ Physiol. 2014;306:H1025–1031. doi: 10.1152/ajpheart.00795.2013

20. Xiao H, Li H, Wang J-J, Zhang J-S, Shen J, An X-B, Zhang C-C, Wu J-M, Song Y, Wang X-Y, et al. IL-18 cleavage triggers cardiac inflammation and fibrosis upon β-adrenergic insult. European Heart Journal. 2017;39:60–69. doi: 10.1093/eurheartj/ehx261

21. O’Brien LC, Mezzaroma E, Van Tassell BW, Marchetti C, Carbone S, Abbate A, Toldo S. Interleukin-18 as a therapeutic target in acute myocardial infarction and heart failure. Mol Med. 2014;20:221–229. doi: 10.2119/molmed.2014.00034

22. Yu Q, Vazquez R, Khojeini EV, Patel C, Venkataramani R, Larson DF. IL-18 induction of osteopontin mediates cardiac fibrosis and diastolic dysfunction in mice. Am J Physiol Heart Circ Physiol. 2009;297:H76–85. doi: 10.1152/ajpheart.01285.2008

23. Yamaoka-Tojo M, Tojo T, Inomata T, Machida Y, Osada K, Izumi T. Circulating levels of interleukin 18 reflect etiologies of heart failure: Th1/Th2 cytokine imbalance exaggerates the pathophysiology of advanced heart failure. J Card Fail. 2002;8:21–27. doi: 10.1054/jcaf.2002.31628

24. Jia X, Buckley L, Sun C, Al Rifai M, Yu B, Nambi V, Virani SS, Selvin E, Matsushita K, Hoogeveen RC, et al. Association of interleukin-6 and interleukin-18 with cardiovascular disease in older adults: Atherosclerosis Risk in Communities study. European Journal of Preventive Cardiology. 2023;30:1731–1740. doi: 10.1093/eurjpc/zwad197

25. Kaptoge S, Seshasai SRK, Gao P, Freitag DF, Butterworth AS, Borglykke A, Di Angelantonio E, Gudnason V, Rumley A, Lowe GDO, et al. Inflammatory cytokines and risk of coronary heart disease: new prospective study and updated meta-analysis. European Heart Journal. 2013;35:578–589. doi: 10.1093/eurheartj/eht367

26. Jefferis BJ, Papacosta O, Owen CG, Wannamethee SG, Humphries SE, Woodward M, Lennon LT, Thomson A, Welsh P, Rumley A, et al. Interleukin 18 and coronary heart disease: prospective study and systematic review. Atherosclerosis. 2011;217:227–233. doi: 10.1016/j.atherosclerosis.2011.03.015

27. Henry A, Gordillo-Marañón M, Finan C, Schmidt AF, Ferreira JP, Karra R, Sundström J, Lind L, Ärnlöv J, Zannad F. Therapeutic targets for heart failure identified using proteomics and Mendelian randomization. Circulation. 2022;145:1205–1217.

28. Minikel EV, Painter JL, Dong CC, Nelson MR. Refining the impact of genetic evidence on clinical success. Nature. 2024;629:624–629. doi: 10.1038/s41586-024-07316-0

29. Gill D, Georgakis MK, Walker VM, Schmidt AF, Gkatzionis A, Freitag DF, Finan C, Hingorani AD, Howson JM, Burgess S. Mendelian randomization for studying the effects of perturbing drug targets. Wellcome open research. 2021;6.

30. Kaplanski G. Interleukin-18: Biological properties and role in disease pathogenesis. Immunological reviews. 2018;281:138–153.

31. Gabay C, Fautrel B, Rech J, Spertini F, Feist E, Kötter I, Hachulla E, Morel J, Schaeverbeke T, Hamidou MA, et al. Open-label, multicentre, dose-escalating phase II clinical trial on the safety and efficacy of tadekinig alfa (IL-18BP) in adult-onset Still’s disease. Annals of the Rheumatic Diseases. 2018;77:840–847. doi: 10.1136/annrheumdis-2017-212608

32. Robertson MJ, Kirkwood JM, Logan TF, Koch KM, Kathman S, Kirby LC, Bell WN, Thurmond LM, Weisenbach J, Dar MM. A dose-escalation study of recombinant human interleukin-18 using two different schedules of administration in patients with cancer. Clinical Cancer Research. 2008;14:3462–3469.

33. Pilling LC, Atkins JL, Bowman K, Jones SE, Tyrrell J, Beaumont RN, Ruth KS, Tuke MA, Yaghootkar H, Wood AR. Human longevity is influenced by many genetic variants: evidence from 75,000 UK Biobank participants. Aging (Albany NY). 2016;8:547.

34. Bouras E, Karhunen V, Gill D, Huang J, Haycock PC, Gunter MJ, Johansson M, Brennan P, Key T, Lewis SJ. Circulating inflammatory cytokines and risk of five cancers: a Mendelian randomization analysis. BMC medicine. 2022;20:1–15.

35. Folkersen L, Gustafsson S, Wang Q, Hansen DH, Hedman ÅK, Schork A, Page K, Zhernakova DV, Wu Y, Peters J. Genomic and drug target evaluation of 90 cardiovascular proteins in 30,931 individuals. Nature metabolism. 2020;2:1135–1148.

36. Richmond RC, Smith GD. Mendelian randomization: concepts and scope. Cold Spring Harbor perspectives in medicine. 2022;12:a040501.

37. Woolf B, Zagkos L, Gill D. TwoStepCisMR: a novel method and R package for attenuating bias in cis-mendelian randomization analyses. Genes. 2022;13:1541.

38. Burgess S, Thompson DJ, Rees JM, Day FR, Perry JR, Ong KK. Dissecting causal pathways using Mendelian randomization with summarized genetic data: application to age at menarche and risk of breast cancer. Genetics. 2017;207:481–487.

39. Zuber V, Grinberg NF, Gill D, Manipur I, Slob EA, Patel A, Wallace C, Burgess S. Combining evidence from Mendelian randomization and colocalization: Review and comparison of approaches. The American Journal of Human Genetics. 2022;109:767–782.

40. Zheng J, Haberland V, Baird D, Walker V, Haycock PC, Hurle MR, Gutteridge A, Erola P, Liu Y, Luo S. Phenome-wide Mendelian randomization mapping the influence of the plasma proteome on complex diseases. Nature genetics. 2020;52:1122–1131.

41. Mezzaroma E, Abbate A, Toldo S. The inflammasome in heart failure. Curr Opin Physiol. 2021;19:105–112. doi: 10.1016/j.cophys.2020.09.013

42. Liu Y, Luo D, Liu E, Liu T, Xu G, Liang X, Yuan M, Zhang Y, Chen X, Chen X. MiRNA21 and IL-18 levels in left atrial blood in patients with atrial fibrillation undergoing cryoablation and their predictive value for recurrence of atrial fibrillation. Journal of Interventional Cardiac Electrophysiology. 2022;64:111–120.

43. Luan Y, Guo Y, Li S, Yu B, Zhu S, Li S, Li N, Tian Z, Peng C, Cheng J. Interleukin-18 among atrial fibrillation patients in the absence of structural heart disease. Europace. 2010;12:1713–1718.

44. Schmidt AF, Bourfiss M, Alasiri A, Puyol-Anton E, Chopade S, van Vugt M, van der Laan SW, Gross C, Clarkson C, Henry A. Druggable proteins influencing cardiac structure and function: Implications for heart failure therapies and cancer cardiotoxicity. Science advances. 2023;9:eadd4984.

45. van Vugt M, Finan C, Chopade S, Providencia R, Bezzina CR, Asselbergs FW, van Setten J, Schmidt AF. Integrating metabolomics and proteomics to identify novel drug targets for heart failure and atrial fibrillation. Genome Medicine. 2024;16:120.

46. Wang Y-H, Fu L, Wang B, Li S-F, Sun Z, Luan Y. Genetic variants of interleukin-18 are associated with reduced risk of atrial fibrillation in a population from Northeast China. Gene. 2017;626:269–274.

47. Savarese G, Becher PM, Lund LH, Seferovic P, Rosano GM, Coats AJ. Global burden of heart failure: a comprehensive and updated review of epidemiology. Cardiovascular research. 2022;118:3272–3287.

48. Collaborators GBDS. Global, regional, and national burden of stroke, 1990-2016: a systematic analysis for the Global Burden of Disease Study 2016. Lancet Neurol. 2019;18:439–458. doi: 10.1016/S1474-4422(19)30034-1

49. Kommu S, Arepally S. The effect of colchicine on atrial fibrillation: a systematic review and meta-analysis. Cureus. 2023;15.

50. Van Tassell BW, Canada J, Carbone S, Trankle C, Buckley L, Oddi Erdle C, Abouzaki NA, Dixon D, Kadariya D, Christopher S. Interleukin-1 blockade in recently decompensated systolic heart failure: results from REDHART (Recently Decompensated Heart Failure Anakinra Response Trial). Circulation: Heart Failure. 2017;10:e004373.

51. Tsimikas S, Gordts PL, Nora C, Yeang C, Witztum JL. Statin therapy increases lipoprotein (a) levels. European heart journal. 2020;41:2275–2284.

52. Schiff MH, Kremer JM, Jahreis A, Vernon E, Isaacs JD, van Vollenhoven RF. Integrated safety in tocilizumab clinical trials. Arthritis Research & Therapy. 2011;13:R141. doi: 10.1186/ar3455

53. Moser JC, Sullivan R, Taylor MH, Puzanov I, Falchook GS, Sznol M, Paton VE, Chonzi D, Garofalo B, Sonnemann M, et al. 736 A phase 1/2 open-label, dose-escalation study of ST-067, a decoy-resistant IL-18 cytokine, given as a monotherapy and with pembrolizumab in advanced solid tumor malignancies. Journal for ImmunoTherapy of Cancer. 2023;11:A829–A829. doi: 10.1136/jitc-2023-SITC2023.0736

54. Wang J, Sun C, Gerdes N, Liu C, Liao M, Liu J, Shi MA, He A, Zhou Y, Sukhova GK, et al. Interleukin 18 function in atherosclerosis is mediated by the interleukin 18 receptor and the Na-Cl co-transporter. Nature Medicine. 2015;21:820–826. doi: 10.1038/nm.3890

