## Supplementary Methods for "Genetically Predicted IL-18 Inhibition and Risk of Cardiovascular Events: A Mendelian Randomization Study"


1. **Extended Introduction**
2. **Extended Methods**
   **2.1. Study Populations**
   **2.2. Statistical Methods**
3. **Strobe MR Checklist**

**Extended Introduction**

Mendelian Randomization (MR) is a method that uses genetic variants as instrumental variables to explore causal relationships between exposures and outcomes. These variants are distributed randomly across the population due to Mendel's laws of segregation and independent assortment, effectively randomizing individuals to different levels of exposure or outcomes. This natural randomization mirrors the process used in randomized control trials, providing an unconfounded estimate of a treatment effect on an outcome. Causal estimates from MR analyses rely on three key assumptions: (i) relevance, where the genetic instrument must be associated with the exposure; (ii) independence, where the instrument is not associated with any confounders of the exposure-outcome relationship; and (iii) exclusion restriction, where the instrument affects the outcome solely through its effect on the exposure. The relevance assumption can typically be confirmed by generating $F$-statistics, with an $F$-statistic above 10 considered sufficient to avoid weak instrument bias. However, the other two assumptions can only be partially addressed. Where possible, multivariable MR can adjust for known confounders statistically. For the exclusion restriction assumption, sensitivity analyses such as MR-Egger regression or the weighted median approach can help identify and correct for any potential pleiotropic effects, where the genetic variants influence the outcome through pathways other than through the exposure of interest. Recently, the application of Mendelian Randomization (MR) has expanded to include the investigation of drug targets. Most drugs target specific proteins, and genetic variants around the gene loci of these proteins can assess the impact of altering the drug target protein on relevant outcomes. Such variants, known as cis-variants, are located within or near the gene locus encoding the drug target protein and are used in MR to explore both existing and novel drug targets. Drug target MR adheres to the same foundational assumptions as traditional MR but includes additional validation steps to confirm that the genetic instruments reflect the pharmacological effects of interest.

### Extended Methods

### Study Populations

#### *Interleukin 18*

Summary statistics for circulating Interleukin-18 concentration were obtained from the Scallop Consortium's genome-wide association meta-analysis of 90 OLINK-measured proteins from 13 contributing cohorts containing 21,758 European individuals.[1] Each cohort measured levels of circulating IL-18 using the OLINK IL-18 assay and obtained genetic associations with an additive effect model adjusted for population stratification and study-specific covariates. Further information on all summary statistics used in this study is available in Supplementary Table 1.

#### *Stroke*

Summary statistics for ischaemic stroke and its three major subtypes (large artery, small-vessel, and cardioembolic) were obtained from GIGASTROKE.[2] GIGASTROKE is a large cross-ancestry genome-wide association meta-analysis with 110,182 stroke cases and 1,503,898 controls in European ancestry. Associations between genotypes and four stroke phenotypes (all-ischemic stroke, cardioembolic stroke, large artery stroke, and small vessel stroke) were analyzed using an additive effects model. Adjustments were made for age, sex, principal components of population stratification, and study-specific covariates.

#### *Coronary artery disease*

Summary statistics for coronary artery disease (CAD) were derived from Cardiogram, a large genome-wide association study (GWAS) that included 181,522 cases among 1,165,690 participants of predominantly European ancestry.[3] Additive logistic models (or logistic mixed models) were used to determine variant-outcome associations adjusting for study-specific covariates and potential ancestry effects.

#### *Atrial fibrillation*

Summary statistics for atrial fibrillation (AF) were obtained from Nielsen et al.'s GWAS, which included 60,620 clinically diagnosed AF cases and 970,216 controls.[4] For primary analyses, we used a subset of this dataset that excluded the UK Biobank (UKB), consisting of 45,800 cases and 589,297 controls.[4] The full dataset, including UKB included, was used for mediation analyses. In our replication analyses, we utilized summary statistics for European ancestry AF cases from the Pan-UKB GWAS, which included 19,218 cases and 401,313 controls.

#### *Heart failure*

Summary statistics for all-cause and non-ischemic heart failure were obtained from Aragam et al.'s GWAS in the UK Biobank (UKB), which included 6,504 cases and 387,652 controls.[5] Non-ischemic heart failure was defined as heart failure diagnosed in UKB individuals who exhibited left ventricular dysfunction and did not have diagnosed CAD. For replication analysis, we used a GWAS of heart failure encompassing 10,976 cases and 437,573 controls from five US-based cohorts.[6]

***Dilated cardiomyopathy***

Summary statistics for dilated cardiomyopathy were obtained from Ning et al.'s GWAS in UKB, which included 1303 cases and 5281 controls.[7] Additive logistic models were used to obtain variant-outcome associations and adjusted for covariates including age, sex, BMI, smoking status, alcohol intake frequency, and the first ten principal components.

***Aortic stenosis and conduction disorders***

Summary statistics for calcific aortic stenosis that led to operative replacement (9870 cases, 402311 controls), left bundle branch block (2130 cases, 312154 controls), right bundle branch block (1046 cases, 312154 controls) and atrioventricular block (6097 cases, 312154 controls) were obtained from FinnGenn (data release 10).[8]

***Peripheral arterial disease***

Summary statistics for peripheral arterial disease were obtained from Van Zuydam et al.'s genome-wide meta-analysis comprising 11 independent cohorts of individuals of European ancestry, which included 12,086 cases and 449,548 controls.[9]

***Abdominal aortic aneurysm***

Summary statistics for abdominal aortic aneurysm were obtained from the Global Biobank Meta-analysis of 8,163 cases and 1,256,755 controls in individuals of European ancestry from 23 international biobanks.

***Cardiac magnetic resonance imaging traits***

Summary statistics for cardiac magnetic resonance imaging traits were obtained from four GWAS conducted in UKB. Thanaj et al. calculated left atrial volume in 39,559 individuals using segmented images indexed to body surface area. Aung et al. measured left ventricular traits in 16,920 individuals free from prevalent heart failure or myocardial infarction, using manual annotation for a subset of 5,000 individuals and a fully convolutional neural network trained for the remaining CMR studies.[10] Aung et al. used a similar combination of manual segmentation and automatic annotation with a deep learning algorithm to measure right ventricular traits in 29,506 individuals without prevalent heart failure or myocardial infarction.[10] Nauffal et al. used a machine learning model to measure native myocardial T1 time, a marker of myocardial fibrosis, in 41,505 UK Biobank participants.[11]

***Biomarkers and longevity***

Summary statistics for lipid traits in European ancestry individuals were derived from the Global Lipids Genetics Consortium (N = 1,320,016) and a UKB GWAS conducted by Richardson et al (N = 115,082). For inflammatory markers, summary statistics were sourced as follows: CRP from Said et al.'s GWAS involving over 500,000 individuals; tumor necrosis factor from The INTERVAL Study (N = 3,301); interferon-gamma from Ahola-Olli et al.'s GWAS (N = 7,701); IL-1 beta from deCODE (N = 35,559); CXCL10 from Gudjonsson et al.'s GWAS (N = 5,366), and IL-6 from a genome-wide meta-analysis by Zhao et al (N = 14,743). Summary statistics for body mass index ((N = 461,460) and systolic and diastolic blood pressure (N = 436,419) were obtained from the Integrative Epidemiology Unit (IEU) OpenGWAS project in UKB individuals of European ancestry. Summary statistics for longevity were derived from Timmers et al.'s genome-wide association study (GWAS) of parental attained age, based on data from 415,311 fathers and 412,937 mothers of European descent participants in the UK Biobank (UKB), where the age of parents (in years) was reported by their offspring.[12]

#### *Cancer*

Summary statistics for lung cancer were obtained from McKay et al.'s GWAS in European ancestry with 29,266 cases and 56,450 controls,[13] while breast cancer from Michailidou et al.'s genome-wide meta-analysis of 46,785 cases and 42,892 controls. Summary statistics for prostate cancer were derived from Wang et al.'s GWAS containing 122,188 cases and 604,640 controls,[14] and colorectal cancer from a meta-analysis of FinnGen and UKB (7,902 cases, 730,965 controls).[15]

#### *Autoimmune and neurological diseases*

Summary statistics for atopic dermatitis (26,954 cases, 768,943 controls), psoriasis (20,788 cases, 806,585 controls), childhood asthma (37,772 cases, 599,390 controls), ulcerative colitis (8,908 cases, 822,951 controls), Crohn's disease (3,721 cases, 828,794 controls), migraine (26,901 cases, 707,900 controls) and Parkinson's disease (6,341 cases, 810,749 controls) were obtained from a meta-analysis of FinnGen and UKB.[15] Summary statistics for rheumatoid arthritis were obtained Ishigaki et al.'s GWAS containing 22,350 cases and 74,823 controls of European ancestry,[16] while summary statistics for multiple sclerosis were obtained from Andlauer et al.'s GWAS in a German population (4,888 cases, 10,395 controls).[17]

#### *Infections*

Summary statistics for sepsis requiring hospital admission (11,643 cases, 47,481 controls), urinary tract infection (21,958 cases, 486,484 controls) and cellulitis (12,196 cases, 474,288 controls) were obtained from Hamilton et al.'s GWAS in UKB.[18] Summary statistics for pneumonia were obtained from Sakaue et al.'s meta-analysis of UKB and FinnGen,[19] while summary statistics for tuberculosis were obtained from FinnGen (release 10).[8]

### Statistical methods

#### *Bidirectional MR*

Bidirectional MR is a method that assesses potential reciprocal causal relationships between two traits using genetic variants as instrumental variables.[20] This approach helps to establish whether associations are causal or correlational, and provides insights into the dynamics between complex traits. In our study, we created genetic instruments for each cardiometabolic trait using variants from across the genome that were associated with each trait at genome-wide significance. We performed clumping with a linkage disequilibrium (LD) threshold of $r^2 < 0.001$ and a distance threshold of 10,000 kb, using data from the 1000 Genomes Project. For our heart failure (HF) instrument, due to limited power, we relaxed our significance threshold to $p < 5 \times 10^{-7}$. We then applied two-sample MR to evaluate the causal impact of genetic liability to each cardiometabolic trait on genetically predicted levels of circulating IL-18.

#### *LD-check*

LD Check, introduced by Zheng et al., is an alternative colocalization approach when traditional Bayesian 'coloc' analyses are not well-powered due to a limited number of SNPs.[21] This method involves checking that at least one of the top 30 cis-acting variants in the outcome's summary statistics exhibits strong linkage disequilibrium (LD) with the putative causal variant from the exposure summary statistics ($r^2 > 0.8$). This degree of multicollinearity suggests that the variants cannot be modeled as distinct variables in a regression analysis, implying a shared underlying causal relationship.

### Supplementary references


1. Folkersen L, Gustafsson S, Wang Q, Hansen DH, Hedman ÅK, Schork A, et al. Genomic and drug target evaluation of 90 cardiovascular proteins in 30,931 individuals. *Nature metabolism*. 2020;2:1135-1148
2. Mishra A, Malik R, Hachiya T, Jürgenson T, Namba S, Posner DC, et al. Stroke genetics informs drug discovery and risk prediction across ancestries. *Nature*. 2022;611:115-123
3. Aragam KG, Jiang T, Goel A, Kanoni S, Wolford BN, Atri DS, et al. Discovery and systematic characterization of risk variants and genes for coronary artery disease in over a million participants. *Nature genetics*. 2022;54:1803-1815
4. Nielsen JB, Thorolfsdottir RB, Fritsche LG, Zhou W, Skov MW, Graham SE, et al. Biobank-driven genomic discovery yields new insight into atrial fibrillation biology. *Nature genetics*. 2018;50:1234-1239
5. Aragam KG, Chaffin M, Levinson RT, McDermott G, Choi SH, Shoemaker MB, et al. Phenotypic refinement of heart failure in a national biobank facilitates genetic discovery. *Circulation*. 2019;139:489-501
6. Arvanitis M, Tampakakis E, Zhang Y, Wang W, Auton A, Dutta D, et al. Genome-wide association and multi-omic analyses reveal actn2 as a gene linked to heart failure. *Nature communications*. 2020;11:1122
7. Ning C, Fan L, Jin M, Wang W, Hu Z, Cai Y, et al. Genome-wide association analysis of left ventricular imaging-derived phenotypes identifies 72 risk loci and yields genetic insights into hypertrophic cardiomyopathy. *Nature Communications*. 2023;14:7900

8. Kurki MI, Karjalainen J, Palta P, Sipilä TP, Kristiansson K, Donner KM, et al. Finngen provides genetic insights from a well-phenotyped isolated population. *Nature*. 2023;613:508-518

9. Van Zuydam NR, Stiby A, Abdalla M, Austin E, Dahlström EH, McLachlan S, et al. Genome-wide association study of peripheral artery disease. *Circulation: Genomic and Precision Medicine*. 2021;14:e002862

10. Aung N, Vargas JD, Yang C, Cabrera CP, Warren HR, Fung K, et al. Genome-wide analysis of left ventricular image-derived phenotypes identifies fourteen loci associated with cardiac morphogenesis and heart failure development. *Circulation*. 2019;140:1318-1330

11. Nauffal V, Di Achille P, Klarqvist MDR, Cunningham JW, Hill MC, Pirruccello JP, et al. Genetics of myocardial interstitial fibrosis in the human heart and association with disease. *Nature Genetics*. 2023;55:777-786

12. Timmers PR, Mounier N, Lall K, Fischer K, Ning Z, Feng X, et al. Genomics of 1 million parent lifespans implicates novel pathways and common diseases and distinguishes survival chances. *elife*. 2019;8:e39856

13. McKay JD, Hung RJ, Han Y, Zong X, Carreras-Torres R, Christiani DC, et al. Large-scale association analysis identifies new lung cancer susceptibility loci and heterogeneity in genetic susceptibility across histological subtypes. *Nature genetics*. 2017;49:1126-1132

14. Wang Y-H, Fu L, Wang B, Li S-F, Sun Z, Luan Y. Genetic variants of interleukin-18 are associated with reduced risk of atrial fibrillation in a population from northeast china. *Gene*. 2017;626:269-274

15. Sun BB, Kurki MI, Foley CN, Mechakra A, Chen C-Y, Marshall E, et al. Genetic associations of protein-coding variants in human disease. *Nature*. 2022;603:95-102

16. Ishigaki K, Sakaue S, Terao C, Luo Y, Sonehara K, Yamaguchi K, et al. Multi-ancestry genome-wide association analyses identify novel genetic mechanisms in rheumatoid arthritis. *Nat Genet*. 2022;54:1640-1651

17. Andlauer TF, Buck D, Antony G, Bayas A, Bechmann L, Berthele A, et al. Novel multiple sclerosis susceptibility loci implicated in epigenetic regulation. *Sci Adv*. 2016;2:e1501678

18. Hamilton FW, Thomas M, Arnold D, Palmer T, Moran E, Mentzer AJ, et al. Therapeutic potential of il6r blockade for the treatment of sepsis and sepsis-related death: A mendelian randomisation study. *PLoS Med*. 2023;20:e1004174

19. Sakaue S, Kanai M, Tanigawa Y, Karjalainen J, Kurki M, Koshiba S, et al. A cross-population atlas of genetic associations for 220 human phenotypes. *Nature Genetics*. 2021;53:1415-1424

20. Richmond RC, Smith GD. Mendelian randomization: Concepts and scope. *Cold Spring Harbor perspectives in medicine*. 2022;12:a040501

21. Zheng J, Haberland V, Baird D, Walker V, Haycock PC, Hurle MR, et al. Phenome-wide mendelian randomization mapping the influence of the plasma proteome on complex diseases. *Nature genetics*. 2020;52:1122-1131

### STROBE-MR Checklist

| Item No. | Section | | Checklist item | Page No. | Relevant text from manuscript |
|---|---|---|---|---|---|
| 1 | **TITLE and ABSTRACT** | | Indicate Mendelian randomization (MR) as the study's design in the title and/or the abstract if that is a main purpose of the study | 1, 3 | Inserted title and abstract |
| | **INTRODUCTION** | | | | |
| 2 | **Background** | | Explain the scientific background and rationale for the reported study. What is the exposure? Is a potential causal relationship between exposure and outcome plausible? Justify why MR is a helpful method to address the study question | 7-8 | Introduction |
| 3 | **Objectives** | | State specific objectives clearly, including pre-specified causal hypotheses (if any). State that MR is a method that, under specific assumptions, intends to estimate causal effects | 8, 11 | Methods |
| | **METHODS** | | | | |
| 4 | **Study design and data sources** | | Present key elements of the study design early in the article. Consider including a table listing sources of data for all phases of the study. For each data source contributing to the analysis, describe the following: | | |
| | | a) | Setting: Describe the study design and the underlying population, if possible. Describe the setting, locations, and relevant dates, including periods of recruitment, exposure, follow-up, and data collection, when available. | 8-10 | Study Design and Supplementary Methods |
| | | b) | Participants: Give the eligibility criteria, and the sources and methods of selection of participants. Report the sample size, and whether any power or sample size calculations were carried out prior to the main analysis | 9-10 | Data Sources |
| | | c) | Describe measurement, quality control and selection of genetic variants | 10-11 | Instrument variable selection |
| | | d) | For each exposure, outcome, and other relevant variables, describe methods of assessment and diagnostic criteria for diseases | 9-10 | Data sources, Instrumental variable selection, Supplementary methods |

| | | | | | |
|---|---|---|---|---|---|
| | | e) | Provide details of ethics committee approval and participant informed consent, if relevant | 13 | Standard Protocol Approvals, Registrations, and Patient Consents |
| 5 | **Assumptions** | | Explicitly state the three core IV assumptions for the main analysis (relevance, independence and exclusion restriction) as well assumptions for any additional or sensitivity analysis | 11 | Mendelian randomization analyses |
| 6 | **Statistical methods: main analysis** | | Describe statistical methods and statistics used | | |
| | | a) | Describe how quantitative variables were handled in the analyses (i.e., scale, units, model) | 10-13 | Methods |
| | | b) | Describe how genetic variants were handled in the analyses and, if applicable, how their weights were selected | 10-11 | Instrument variable selection |
| | | c) | Describe the MR estimator (e.g. two-stage least squares, Wald ratio) and related statistics. Detail the included covariates and, in case of two-sample MR, whether the same covariate set was used for adjustment in the two samples | 11 | Instrument variable selection |
| | | d) | Explain how missing data were addressed | NA | NA |
| | | e) | If applicable, indicate how multiple testing was addressed | 13 | Reporting and packages |
| 7 | **Assessment of assumptions** | | Describe any methods or prior knowledge used to assess the assumptions or justify their validity | 11 | Mendelian randomization analyses |
| 8 | **Sensitivity analyses and additional analyses** | | Describe any sensitivity analyses or additional analyses performed (e.g. comparison of effect estimates from different approaches, independent replication, bias analytic techniques, validation of instruments, simulations) | 8-12 | Methods |
| 9 | **Software and pre-registration** | | | | |
| | | a) | Name statistical software and package(s), including version and settings used | 13 | Reporting and packages |

| | | | | | |
|---|---|---|---|---|---|
| | | b) | State whether the study protocol and details were pre-registered (as well as when and where) | NA | NA |
| | **RESULTS** | | | | |
| 10 | **Descriptive data** | | | | |
| | | a) | Report the numbers of individuals at each stage of included studies and reasons for exclusion. Consider use of a flow diagram | 10 | Data sources |
| | | b) | Report summary statistics for phenotypic exposure(s), outcome(s), and other relevant variables (e.g. means, SDs, proportions) | 13 | Reporting and packages |
| | | c) | If the data sources include meta-analyses of previous studies, provide the assessments of heterogeneity across these studies | NA | |
| | | d) | For two-sample MR: | | |
| | | | i. Provide justification of the similarity of the genetic variant-exposure associations between the exposure and outcome samples | NA | |
| | | | ii. Provide information on the number of individuals who overlap between the exposure and outcome studies | 9-10 | Methods |
| 11 | **Main results** | | | | |
| | | a) | Report the associations between genetic variant and exposure, and between genetic variant and outcome, preferably on an interpretable scale | Supplementary Tables | Supplementary Table 2 |
| | | b) | Report MR estimates of the relationship between exposure and outcome, and the measures of uncertainty from the MR analysis, on an interpretable scale, such as odds ratio or relative risk per SD difference | 14-18 | Results |
| | | c) | If relevant, consider translating estimates of relative risk into absolute risk for a meaningful time period | N/A | |
| | | d) | Consider plots to visualize results (e.g. forest plot, scatterplot of associations between genetic variants and outcome versus between genetic variants and exposure) | Figure 2-5 | |

| | | | | | |
|---|---|---|---|---|---|
| 12 | **Assessment of assumptions** | | | | |
| | | a) | Report the assessment of the validity of the assumptions | 18 | Results |
| | | b) | Report any additional statistics (e.g., assessments of heterogeneity across genetic variants, such as $I^2$, Q statistic or E-value) | 18 | Results |
| 13 | **Sensitivity analyses and additional analyses** | | | | |
| | | a) | Report any sensitivity analyses to assess the robustness of the main results to violations of the assumptions | 17-18 | Results |
| | | b) | Report results from other sensitivity analyses or additional analyses | 16-18 | Results |
| | | c) | Report any assessment of direction of causal relationship (e.g., bidirectional MR) | 17 | Results |
| | | d) | When relevant, report and compare with estimates from non-MR analyses | 18-22 | Results |
| | | e) | Consider additional plots to visualize results (e.g., leave-one-out analyses) | NA | NA |
| | **DISCUSSION** | | | | |
| 14 | **Key results** | | Summarize key results with reference to study objectives | 18 | Discussion |
| 15 | **Limitations** | | Discuss limitations of the study, taking into account the validity of the IV assumptions, other sources of potential bias, and imprecision. Discuss both direction and magnitude of any potential bias and any efforts to address them | 22-23 | Limitations |
| 16 | **Interpretation** | | | | |
| | | a) | Meaning: Give a cautious overall interpretation of results in the context of their limitations and in comparison with other studies | 18-23 | Discussion |
| | | b) | Mechanism: Discuss underlying biological mechanisms that could drive a potential causal relationship between the investigated exposure and the outcome, and whether the gene-environment equivalence assumption is | 18-13 | Discussion |

| | | | | |
|---|---|---|---|---|
| | | reasonable. Use causal language carefully, clarifying that IV estimates may provide causal effects only under certain assumptions | | |
| | c) | Clinical relevance: Discuss whether the results have clinical or public policy relevance, and to what extent they inform effect sizes of possible interventions | 18-23 | Discussion |
| 17 | **Generalizability** | Discuss the generalizability of the study results (a) to other populations, (b) across other exposure periods/timings, and (c) across other levels of exposure | 22-23 | Limitations |
| | **OTHER INFORMATION** | | | |
| 18 | **Funding** | Describe sources of funding and the role of funders in the present study and, if applicable, sources of funding for the databases and original study or studies on which the present study is based | 23 | |
| 19 | **Data and data sharing** | Provide the data used to perform all analyses or report where and how the data can be accessed, and reference these sources in the article. Provide the statistical code needed to reproduce the results in the article, or report whether the code is publicly accessible and if so, where | 13-14 | Data availability, Supplementary Table 1 |
| 20 | **Conflicts of Interest** | All authors should declare all potential conflicts of interest | 23 | Disclosures |
